# Control Strategies for the Third wave of COVID-19 infection in India: A Mathematical Model Incorporating Vaccine Effectiveness

**DOI:** 10.1101/2021.12.27.21268464

**Authors:** Namitha A Sivadas, Pooja Panda, Ashutosh Mahajan

## Abstract

The waning effectiveness of the COVID-19 vaccines and the emergence of a new variant Omicron has given rise to the possibility of another outbreak of the infection in India. COVID-19 has caused more than 34 million reported cases and 475 thousand deaths in India so far, and it has affected the country at the root level, socially as well as economically. After going through different control measures, mass vaccination has been achieved to a large extent for the highly populous country, and currently under progress. India has already been hit by a massive second wave of infection in April-June, 2021 mainly due to the delta variant, and might see a third wave in the near future that needs to be controlled with effective control strategies. In this paper, we present a compartmental epidemiological model with vaccinations incorporating the dose-dependent effectiveness. We study a possible sudden outbreak of SARS-CoV2 variants in the future, and bring out the associated predictions for various vaccination rates and point out optimum control measures. Our results show that for transmission rate 30% higher than the current rate due to emergence of new variant or relaxation of social distancing conditions, daily new cases can peak to 250k in March 2022, taking the second dose effectiveness dropping to 50% in the future. Combination of vaccination and controlled lockdown or social distancing is the key to tackling the current situation and for the coming few months. Our simulation results show that social distancing measures show better control over the disease spread than the higher vaccination rates.

## I. Introduction

Mutations in SARS-CoV-2 can create variants that have more transmissibility and can be more harmful [1]. Recently, a highly contagious new variant of SARS-CoV-2 Omicron has emerged, the first case of which was reported in Southern African country Botswana, on Nov 11, 2021, and a total of 101 Omicron cases have been reported in India so far. The emergence of the variants like alpha, beta, and delta SARS-CoV-2 was linked with new waves of infections which were seen in many countries [2]. Apart from the emergence of new variants like Omicron, the waning effectiveness of the vaccines is also another cause of concern. It has been observed that even after taking both doses of vaccination, new infected cases have been found [3]. The effectiveness of vaccine Pfizer–BioNTech (BNT162b2) is reported to be dropping to 47% after five months of the second dose of vaccination from the initial 90% after one month of vaccination [4]. In Israel, breakthrough infections in healthcare workers were identified at the hospital, and out of 1497 fully vaccinated workers, 39 were found symptomatic infected [5].

This pandemic has triggered various strategies like visitor restrictions, careful contact limitations for patients having respiratory symptoms and universal masking and many other strategies [6], [7], [8]. Although a number of reported cases reduced with the help of those interventions [9], people had to compromise with their socio-economic loss and lifestyle [10]. COVID 19 control measures in India have come a long way from 21 days lockdown to the establishment of vaccine platforms. The impact of the programs depends upon several factors like supply, optimal strategies to implement, and eligibility to receive vaccine [11]. According to the World Health Organization, 133 COVID-19 vaccines were in the process of development during 2020 [12].Four vaccines that came into account in March 2021 after being permitted on all grounds by Italian and European medicine agencies are named Mod-erna(mRNA-1273), Pfizer-BioNTech(BNT162b2), Johnson and Johnson(JNJ-78436735) Ad26.COV2 and Oxford–straZenecaAZD1222 [13]. At present, AstraZeneca “Covishield”(AZD1222 (ChAdOx1)) and Bharat Biotech “Covaxin”(BBV152) are the vaccines available in India [14].

It is reported that vaccination programs could possibly lessen the health risks of the individuals by nearly 99% in comparison to the no-vaccination situation, and reduced by about 40% in relative to mild Non-pharmaceutical Interventions(NPI) [15]. However, the question of to what extent can we relax NPI while vaccination is going on is still unanswered [15].Yang *et.al*. noticed that the vaccination program without NPIs is too slow to counterbalance the epidemic’s spread and bring down infection load on lives [15]. Only vaccination was unable to control rather a simultaneous action of vaccination, and NPIs were needed to be carried out [1], [16].

Therefore, we need to have a model incorporating vaccinations as well as social distancing that can suggest us optimum control measures for preventing the spread of the disease in the future. One needs to analyze the data of the daily number of infected and vaccinated cases for a longer duration to estimate the sustainability of immunity of vaccines which can put some light on the frequency of vaccine dose whether it needs to be repeated periodically or not [17]. Mathematical modeling is undoubtedly a suitable tool to study the dynamics of infectious diseases and predict their impact in the near future. Different models have been suggested in the literature to combat this threatening disease. Many Extensions of the basic SIR model [18] are available. A mathematical model based on the SEIR model with robust control and non-linear algorithm on variable structure control (VSC) is described in [19]. Compartmental models have been applied for COVID-19 spread in India [20] [21] [22] and an extended SEIR model is elaborated for future analysis [23]. A variational mathematical model to analyze the impact of COVID-19 on the healthcare systems is proposed in [24]. A disease transmission model called SEAMHQRD-V is used to explain the effect of lockdown timing in a better way [25]. A compartmental model reported by Senapati *et.al*. to culminate this disease by applying different phases of interventions [26].

In this work, we extend our model SIPHERD [27], [28] incorporating the dose-dependent vaccine effectiveness to estimate the impact of different vaccination rates in controlling the disease, and call it as SIPHERD-V. We obtain the optimized parameter values of the model from the available data to further predict the future in different scenarios such as varying social distancing conditions, the effectiveness of the vaccine for new variants, vaccination rates, and combined effect of all.

## II. Methods

In this work, we develop a compartmental model that incorporates the vaccinations and their effectiveness. The extent by which the mass vaccination rate can control the spread of the disease is modeled by considering the categories of those who received the first and the second dose of the vaccination separately. The effectiveness of the vaccine is incorporated in the parameters *θ*_1_, *θ*_2_.

We utilise the publicly available data sources for COVID-19 [29] [30] for optimizing the model parameters in MATLAB. It also finds the initial values of parameters involved so that prediction for the future can be done. Here we have taken vaccination parameters into account to monitor the effect of vaccination rates on COVID-19 growth. In our model, we have divided the whole population into different sections and named as Susceptible (S), Exposed (E), Symptomatic (I), Purely Asymptomatic (P), Hospitalized or Quarantined (H), Recovered (R) and Deceased (D).

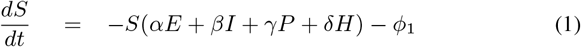

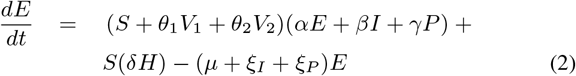

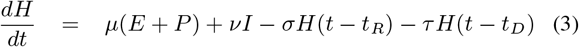

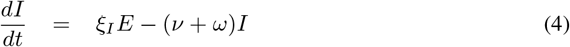

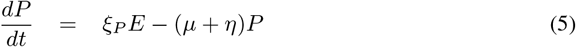

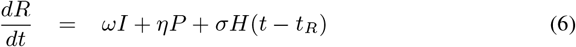

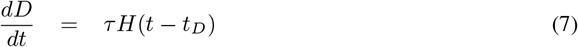

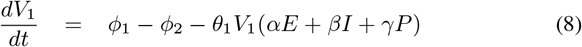

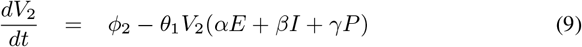

where, *t*_*R*_ and *t*_*D*_ denote the delay related to recovery and death respectively with respect to active cases *H*. The additional entities that we have considered are the fraction of the population that received the first vaccination dose (*V*_1_), second vaccination dose (*V*_2_), and the parameters, the rate of vaccination for the first and second dose as *ϕ*_1_ and *ϕ*_2_ respectively. As the total population in the model is taken constant, we can write

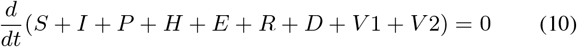

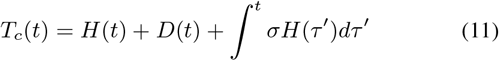

## III. Numerical Implementation and Simulation

The above-coupled equations for the model are solved numerically by dde23 solver routine in MATLAB for ordinary differential equations with a time lag in functions for a set of parameters and initial values. The accurate determination of the parameters that represent the actual situation of the disease spread is essential for determining the future prediction. We take into account the data sets of the total number of confirmed cases, active cases, cumulative deaths, and dose1 and dose2 vaccinations done per day and find the set of parameters that give the solution to the above differential equations close to these actual data sets. The optimized parameters are obtained by finding the minimizer of the standard deviation between the actual data and model values.

For the number of confirmed positive cases, death cases and active cases we collected data from [29], [31], and vaccination per day from [30]. All the parameters that are determined by our model for COVID-19 spread in India are listed in TABLE I. The details of those parameter relating to the characteristics of this disease are already discussed in [28]. The model is optimized for 279 days up to December 4, 2021, to find out the rates of transmissions for the intervals mentioned in TABLE I and other parameter values, and with these parameter values, the model is run for 500 days from March 1, 2021, to July 13, 2022 for obtaining the future predictions.

**Table I:** Parameters values for the SIPHERD-V Model for India

| Parameter | India | Value |
| --- | --- | --- |
| Population | N | 1.38e+09 |
| Starting Date of Simulation | $T_0$ | Mar 1,2021 |
| Number of days till Lockdown | $T_{LD}$ | 20,45,65,90,110,180,250 |
| Transmission rate, E to S before LD | $\alpha_1$ | 0.25 |
| Transmission rate, E to S after LD | $\alpha_2, \alpha_3, \alpha_4, \alpha_5, \alpha_6, \alpha_7$ | 0.20,0.20,0.16,0.13,0.20,0.25,0.23 |
| Transmission rate, I to S before LD | $\beta_1$ | 0.1988 |
| Transmission. rate, I to S after LD | $\beta_2, \beta_3, \beta_4, \beta_5, \beta_6, \beta_7, \beta_8$ | 0.30,0.24,0.13,0.12,0.20,0.24,0.23 |
| Transmission rate, H to S | $\delta$ | 6E-3 |
| Conversion rate, E to I | $\xi_I$ | 0.2 |
| Conversion rate, E to P | $\xi_P$ | 0.072 |
| Detection Probability of E,P | $\mu_0$ | 1.2963 |
| Detection Probability of I | $\nu_0$ | 0.29911 |
| Home Recovery rate of Sym. | $\omega$ | 0.0714 |
| Home Recovery rate of Asy. | $\eta$ | 0.1 |
| Recovery Rate | $\sigma$ | 0.098 |
| Recovery delay | $t_R$ | 5 |
| Mortality delay | $t_D$ | 5 |
| Probability of dose 1 Vaccinated getting infection | $\theta_1$ | 0.4 |
| Probability of dose 2 Vaccinated getting infection | $\theta_2$ | 0.1 |
| Probability of dose 1 Vaccinated getting infection after 4 months | $\theta_1$ | 0.7 |
| Probability of dose 2 Vaccinated getting infection after 4 months | $\theta_2$ | 0.5 |
| Total Initially Infected | $E_0+P_0+I_0$ | 100000 |

## IV. Results and Discussion

We study the effect of vaccinations and social distancing conditions on the disease spread in terms of total cases, active and extinct cases. The effect of new variants of the virus is taken implicitly by changing the transmission rates for the prediction.

### A. Effect of the emergence of highly transmissible variant and relaxed social distancing

The number of daily new cases is showing a slow but steady declining trend in India, however, if a new variant comes with increased transmissibility, or if social distancing conditions are relaxed more than 20% of the current trend, then the cases can rise significantly in the future. In this study, we bring out what would be the effect of the emergence of new variants for a given vaccination rate, and how cautiously the social distancing to be relaxed to keep the daily new cases under control. Schools, colleges, and offices have started opening in India. We consider different scenarios by varying the rate of transmission, which is dependent on both the variant as well as social distancing, and predict the future for this values. If social distancing and lockdown are relaxed, then there can be a surge in infection rates. However, as already the whole country is going through economic losses and this pandemic crisis as well, we need to find a middle ground through which both can be balanced, and can get back to our normal life. From Fig. 2, one can see that the current social distancing can be relaxed by 20% for which daily new cases can rise up to 70k per day around 3 February,2022 and if we relax by 30% then cases can rise up to 244k. The disease evolution has become sensitive towards the rate of transmission since vaccines are showing up waning effectiveness, and even 20% relaxation can lead to 10 million rise in the number of cases around 10 July,2022.

**Fig. 1:**
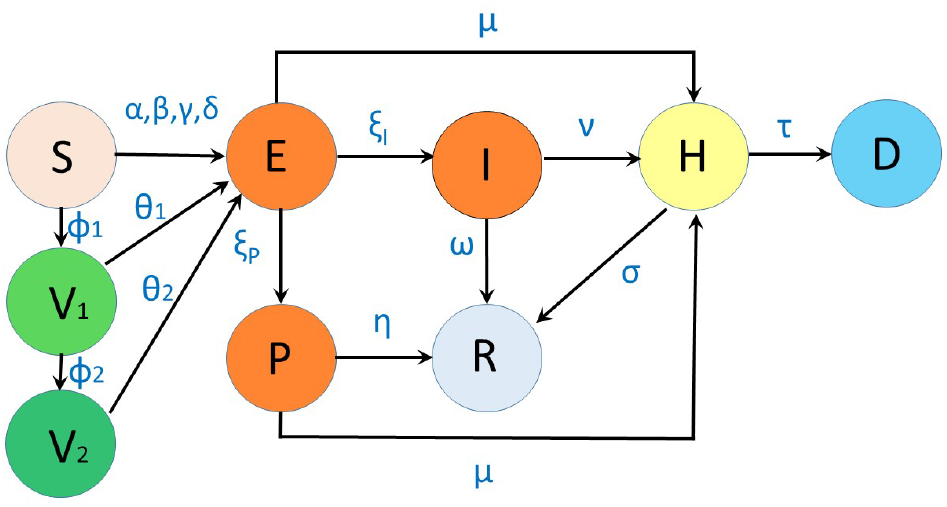
SIPHERD-V model. *α, β, γ, δ* are rates of transmission of infection; *ξ*_*I*_, *ξ*_*P*_ are rates of transfer from exposed to Symptomatic and Asymptomatic; *ω, η, σ* are recovery rates; *µ, ν* are detection rates, and *τ* is mortality rate; *ϕ*_1_, *ϕ*_2_ are the rates of first and second dose of vaccination; *θ*_1_, *θ*_2_ are the rates of vaccinated individuals getting infected.

**Fig. 2:**
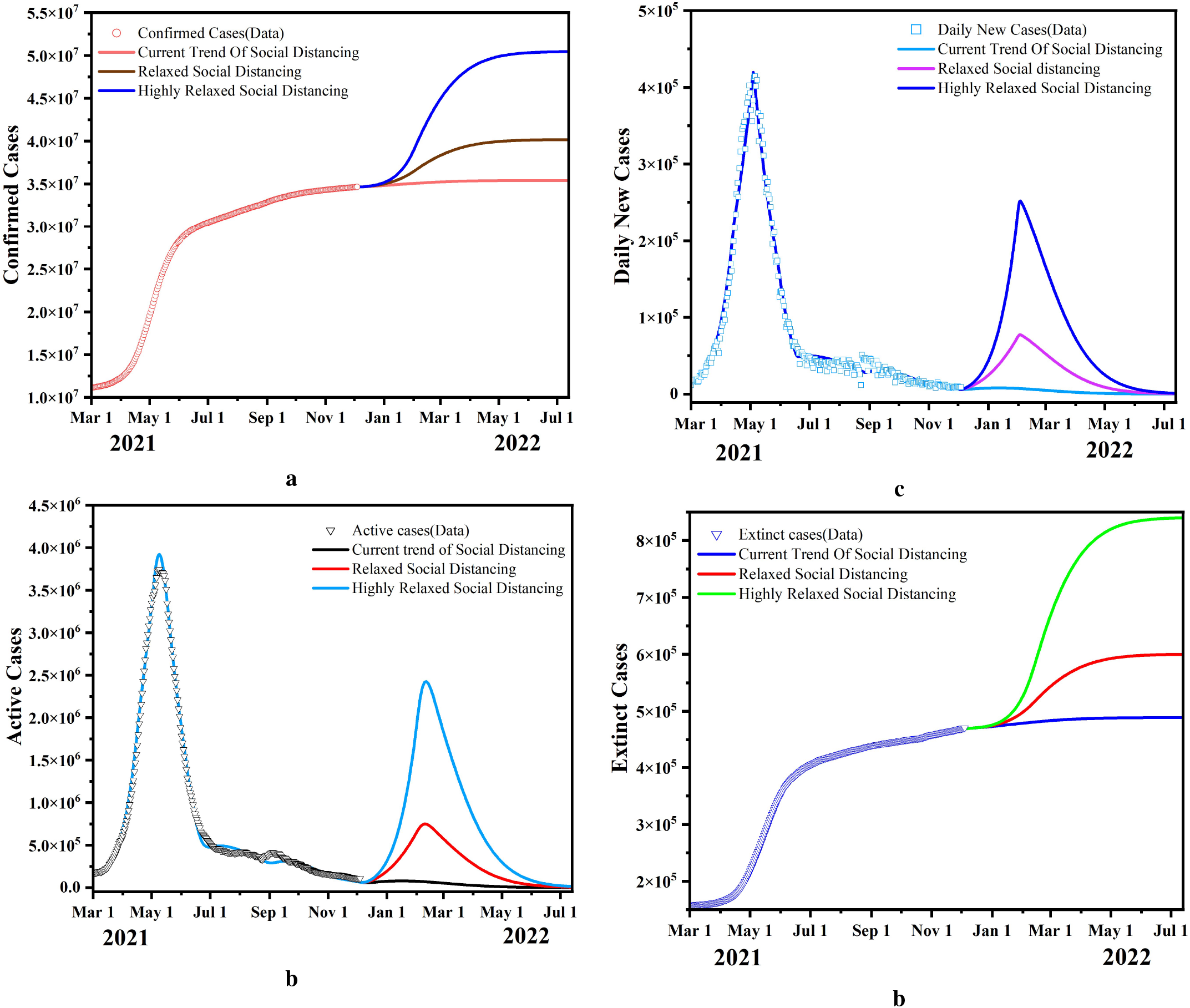
Model predictions for 20% rise in the rate of transmission of the infection. This is taken due to emergence of highly transmissible mutant or the relaxed measures on Social Distancing with vaccination rates continuing at same rate. Model prediction for **a**. the confirmed, daily new cases **b**. active cases **c**. and total deaths **d**. with for moderate and highly relaxed social distancing or highly transmissible variant, and with continuing same current vaccination rate.

### B. Variation in the Effectiveness of Vaccination

Vaccination is obviously effective in stopping the spread of the infection as the fraction of the susceptible people reduces with vaccinations. For the delta variant, the effectiveness of the vaccine was found to be high (93%) after the second dose of vaccination for one month, then it declined to 53 percent after 120 days [4]. In this way, if the immunity fades away with time, the disease is more likely to spread in the future as a fully vaccinated individual would be susceptible to the infection. The probability of getting infected even after vaccination dose1 and dose2 is modeled as *θ*_1_ and *θ*_2_, respectively as seen in Fig. 1, and plays a crucial role in the evolution. Three different cases are compared by varying the effectiveness(1-*θ*) of the doses. Since the effectiveness is reported to decline from 88% to 47% within five months of a vaccine shot [4], we take an “average time effect” value of the effectiveness for the second dose to be 70% and for the first dose to be 50%. For the value of *θ*_1_ and *θ*_2_, 0.5 and 0.3, the number of daily new cases are around 3k, whereas, for [0.7,0.5], [0.8,0.6] it rises to 60k, 268k respectively around 21 february,2021 as seen in Fig. 3.

**Fig. 3:**
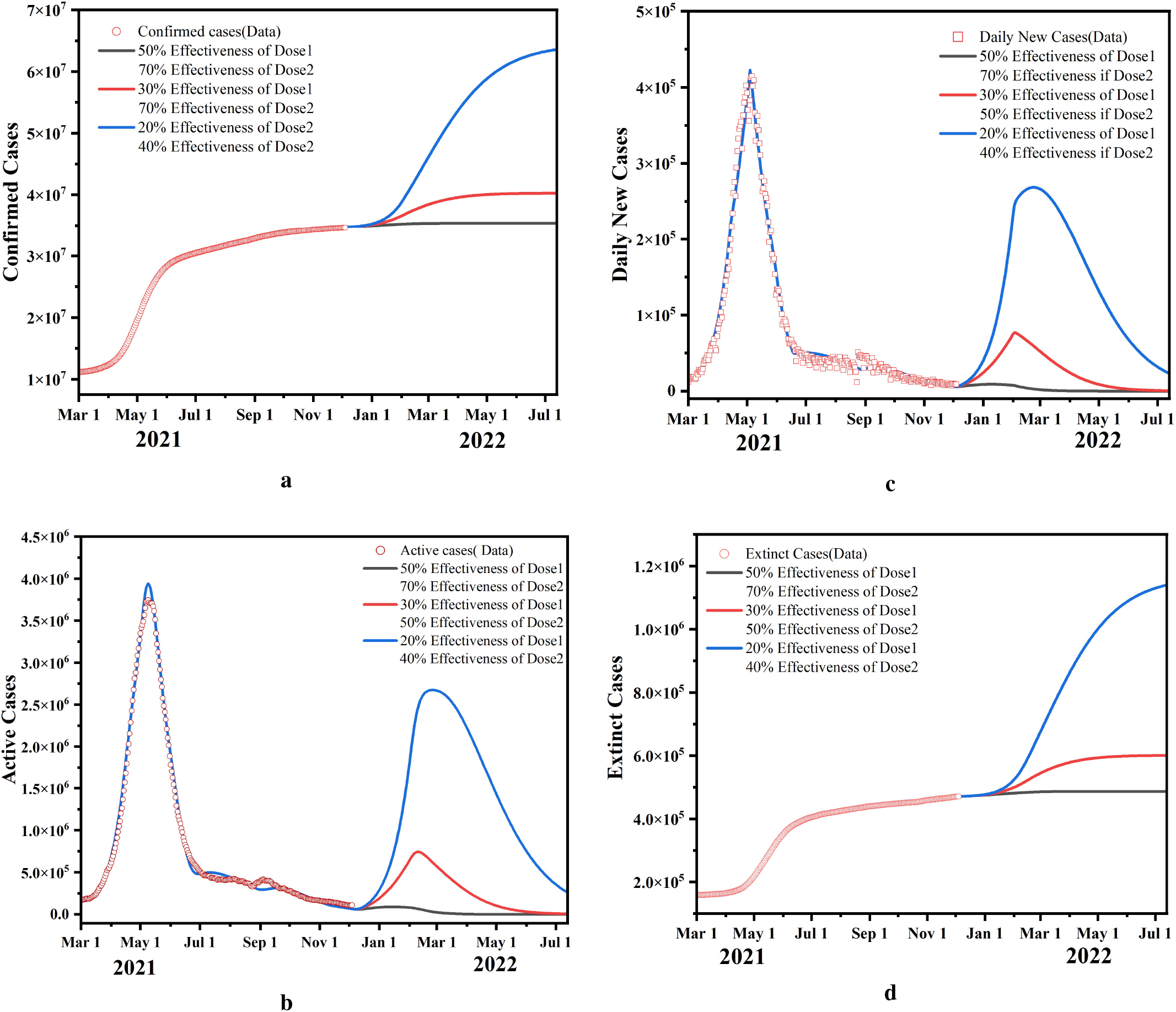
Waning Effectiveness of the Vaccines: Predictions for reduced effectiveness of the vaccines with time or due a future mutation of the virus for which vaccines turn ineffective. The rates of vaccinated people getting infection is taken as the current trend.

### C. Impact of Increase in rate of Vaccination

The daily number of vaccinations for the first and the second dose are the model parameters *ϕ*_1_ and *ϕ*_2_ respectively. How an aggressive increase in the vaccination can control the disease spread is demonstrated in Fig. 5. We consider different vaccination profiles as shown in Fig. 5 c. A total of 9 million, 10 million, and 11 million doses per day are taken on 5th December, 2021 and gradually decreased by 30k per day as the number of people easily available for vaccination is going to show a diminishing trend. To see the extent to which an elevated vaccination rate can control the disease, we take the rate of transmission to be 30% more than the current value. It can be seen in Fig. 5b, the daily new cases peak at 217k for 10 million vaccines on 5th December 2021, and decrease to 189k for 11 million doses. It is observed that on increasing the rate of vaccination by 1 million, a decreasing trend is seen for extinct cases. When the vaccination doses per day increased from 9 to 10 to 11 million doses per day, the extinct cases decreased from 838k to 764k to 710k around 3 July,2022.

**Fig. 4:**
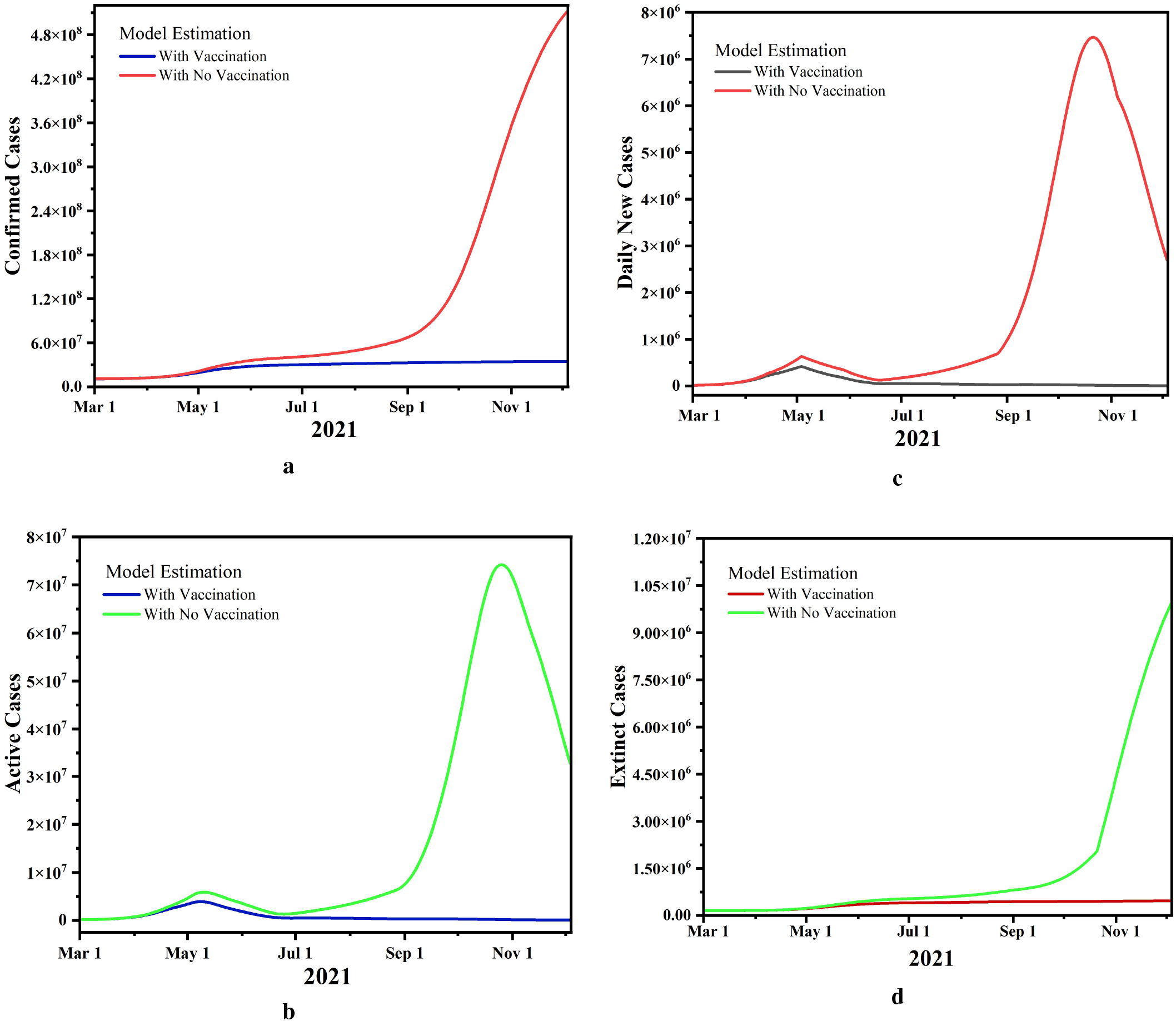
Disease Evolution without any Vaccination Program: Comparison of current evolution of the disease with a situation if no vaccinations were given, and rates of transmission are taken the same with vaccination.

**Fig. 5:**
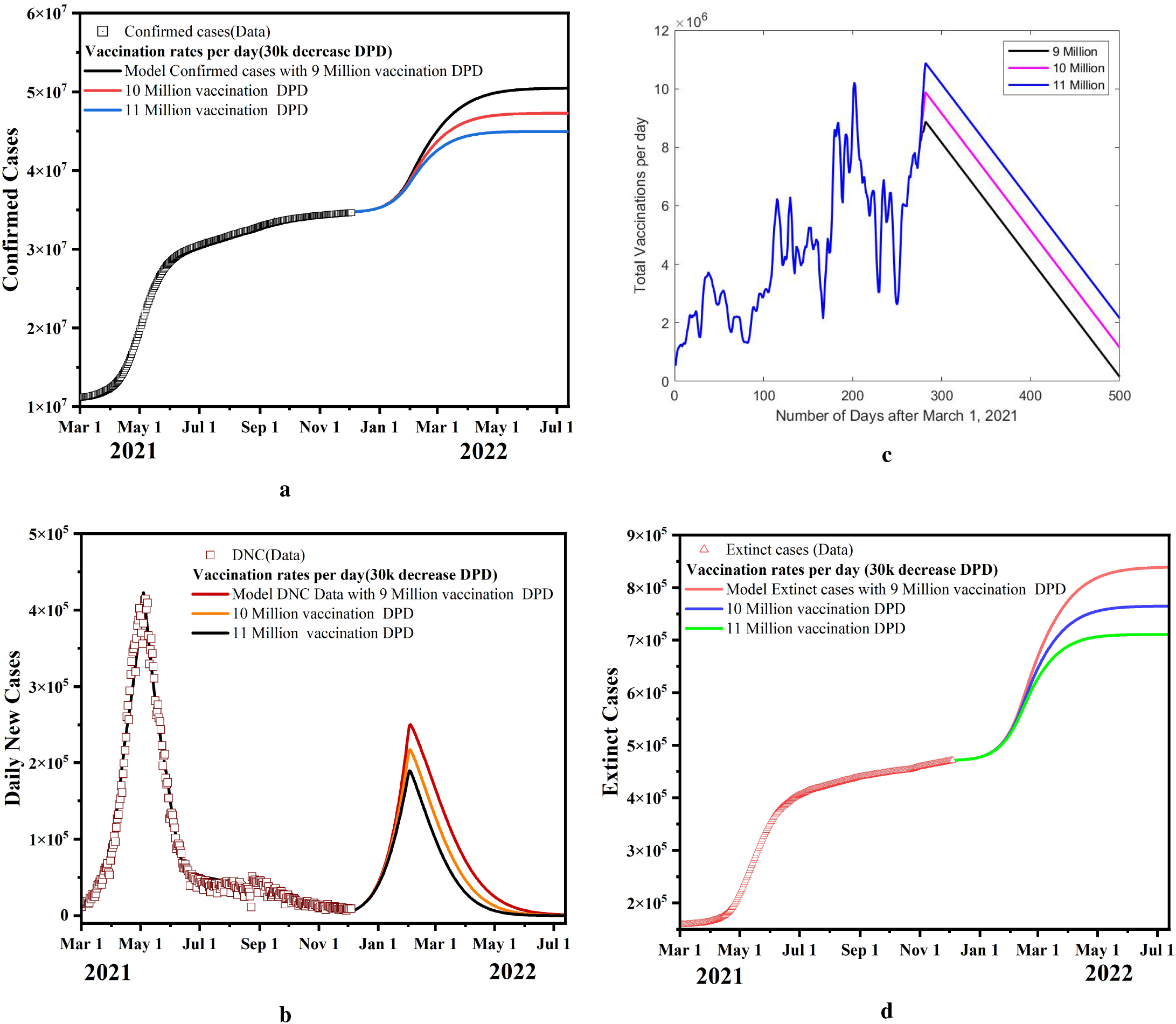
Effect of Increase in vaccination rate: Model predictions for the confirmed, Daily new cases and total deaths for 9 million, 10 million and 11 million vaccinations per day with 30k decrease per day. The total vaccination per day profiles for the 500 days for this study is plotted in b), where the total vaccinations are steadily decremented by 30k per day after December 5, 2021

### D. Comparison of Disease Evolution with and without Vaccination

India is witnessing this dreadful situation even after implementing NPIs and vaccination programs but what would have been the case if the vaccine couldn’t be produced in such a short time and to what extent it would have worsened the situation can be understood from the modeling and simulation. We have done a comparative study between the actual situation and a hypothetical scenario without any vaccination program for the past 279 days keeping the rates of transmission the same for both. Taking out the difference between these two situations, it is found that the number of confirmed cases could have risen to 508 millions from 34.6 millions that indicating the severity of the pandemic in the absence of a vaccine. Our simulation study demonstrated in Fig. 4 shows that the vaccination program has reduced deaths by almost 95%.

### E. Third wave possibility in India and Optimum Control strategies

We study a scenario in which the rate of transmission is taken increased by 30%, and after a month if the control strategies are to be brought in, then what would be the best option for the decision making authorities is investigated. We compared the social distancing impact with the increase in vaccination rates. The vaccination rates are taken elevated to 11 million doses per day. It can be seen from Fig. 6 that in such a situation, an increase in vaccination has a marginal effect as compared to social distancing strictness. After 30 days, the rates of transmission are taken as the current value, and vaccinations are taken 11 million per day with a steady decrease of 30k per day for comparison purpose.

**Fig. 6:**
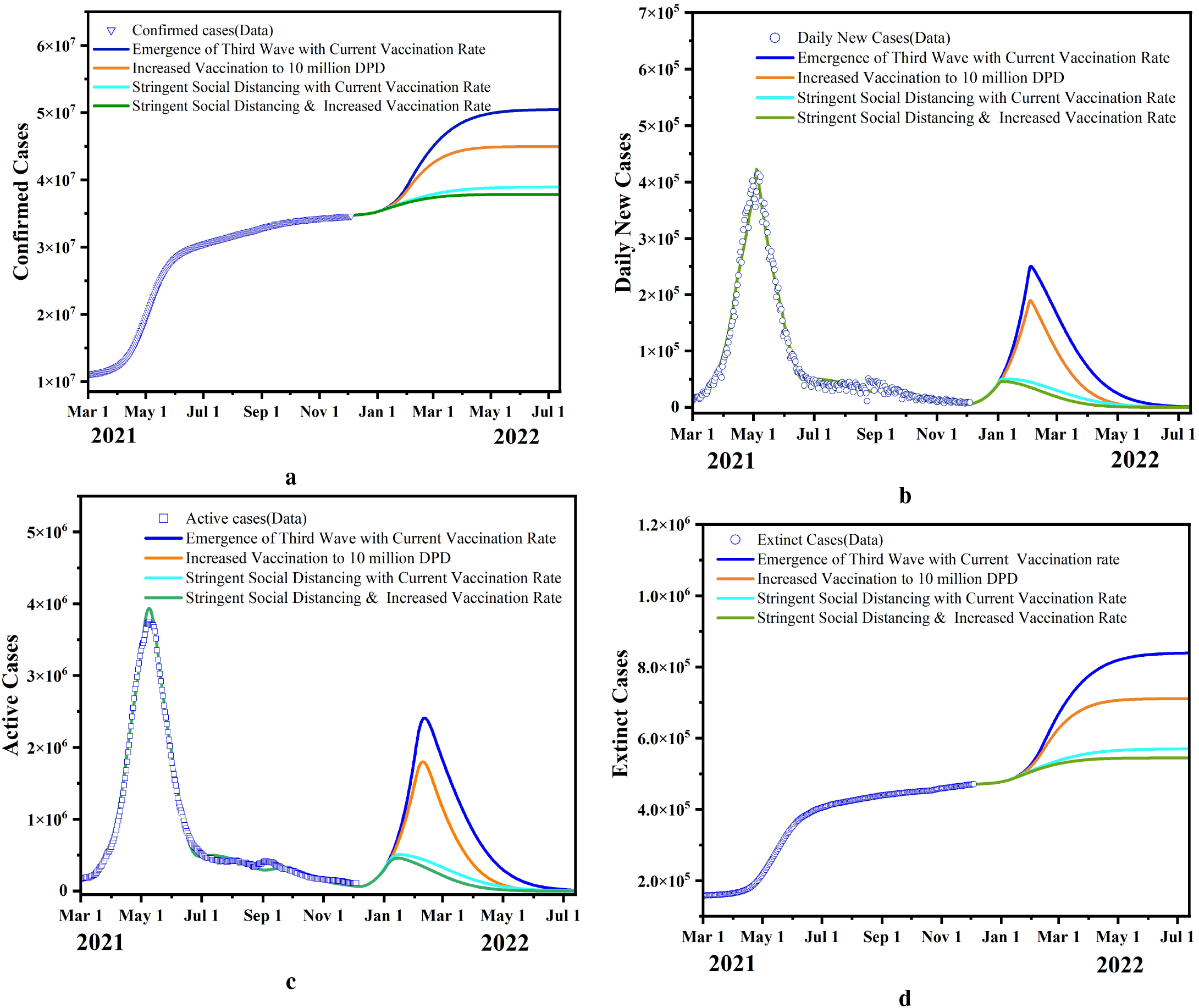
Third wave control measures: Model predictions for the confirmed, active and total deaths and daily new cases for different control measures such as only increase in vaccination, only strict social distancing, and combination of both taken onward 5 Jan 2022.

The mortality rate for the future is taken the same as the current value which could be less for the Omicron as after vaccination, less severe cases requiring hospitalization have been observed.

## V. Conclusion

SIPHERD-V model is applied to analyze the effect of vaccination programs and social distancing on the total confirmed, active, death and daily new cases. We demonstrate how well India’s vaccination program has controlled the COVID-19 situation. The effectiveness of the vaccine is the most critical parameter now for the disease spread and which is waning with time. We show different outcomes based on variation in the effectiveness of the vaccine. Decreasing the value of effectiveness of the fully vaccinated to 50% and the emergence of highly transmissible variant of SARS-CoV-2 imply that the third wave of infection can hit India which can peak around March 2022.

We bring out that in case a variant Omicron spreads significantly after 30-60 days, then social distancing has more impact than the increase in vaccination rate. Different profiles vaccinations with time are investigated and our simulation results show that there can be a steady reduction in a number of active cases and new cases per day after March 2022 if the effective control measures are implemented.

## Data Availability

All data produced are available online at
https://www.worldometers.info/coronavirus.
https://ourworldindata.org/grapher/full-list-covid-19-tests-per-day.
https://www.covid19india.org.

## References

[1] A. Ramos, M. Vela-Pérez, M. Ferrández, A. Kubik, and B. Ivorra, “Modeling the impact of sars-cov-2 variants and vaccines on the spread of covid-19,” Preprint at ResearchGate https://doi.org/10.13140/RG, vol. 2, no. 32580.24967, p. 2, 2021.

[2] A. Fontanet, B. Autran, B. Lina, M. P. Kieny, S. S. A. Karim, and D. Sridhar, “Sars-cov-2 variants and ending the covid-19 pandemic,” The Lancet, vol. 397, no. 10278, pp. 952–954, 2021.

[3] L. J. Abu-Raddad, H. Chemaitelly, and A. A. Butt, “Effectiveness of the bnt162b2 covid-19 vaccine against the b. 1.1. 7 and b. 1.351 variants,” New England Journal of Medicine, 2021.

[4] S. Y. Tartof, J. M. Slezak, H. Fischer, V. Hong, B. K. Ackerson, O. N. Ranasinghe, T. B. Frankland, O. A. Ogun, J. M. Zamparo, S. Gray et al., “Effectiveness of mrna bnt162b2 covid-19 vaccine up to 6 months in a large integrated health system in the usa: a retrospective cohort study,” The Lancet, vol. 398, no. 10309, pp. 1407–1416, 2021.

[5] M. Bergwerk, T. Gonen, Y. Lustig, S. Amit, M. Lipsitch, C. Cohen, M. Mandelboim, E. G. Levin, C. Rubin, V. Indenbaum et al., “Covid-19 breakthrough infections in vaccinated health care workers,” New England Journal of Medicine, vol. 385, no. 16, pp. 1474–1484, 2021.

[6] X. Wang, E. G. Ferro, G. Zhou, D. Hashimoto, and D. L. Bhatt, “Association between universal masking in a health care system and sars-cov-2 positivity among health care workers,” Jama, vol. 324, no. 7, pp. 703–704, 2020.

[7] Y.-A. Liu, Y.-C. Hsu, M.-H. Lin, H.-T. Chang, T.-J. Chen, L.-F. Chou, and S.-J. Hwang, “Hospital visiting policies in the time of coronavirus disease 2019: a nationwide website survey in taiwan,” Journal of the Chinese Medical Association, 2020.

[8] L. E. Wee, E. P. Conceicao, X. Y. J. Sim, K. K. K. Ko, M. L. Ling, and I. Venkatachalam, “Reduction in healthcare-associated respiratory viral infections during a covid-19 outbreak,” Clinical Microbiology and Infection, vol. 26, no. 11, p. 1579, 2020.

[9] V. Priesemann, M. M. Brinkmann, S. Ciesek, S. Cuschieri, T. Czypionka, G. Giordano, D. Gurdasani, C. Hanson, N. Hens, E. Iftekhar et al., “Calling for pan-european commitment for rapid and sustained reduction in sars-cov-2 infections,” The lancet, vol. 397, no. 10269, pp. 92–93, 2021.

[10] J. Zhao, H. Jin, X. Li, J. Jia, C. Zhang, H. Zhao, W. Ma, Z. Wang, Y. He, J. Lee et al., “Disease burden attributable to the first wave of covid-19 in china and the effect of timing on the cost-effectiveness of movement restriction policies,” Value in Health, vol. 24, no. 5, pp. 615–624, 2021.

[11] W. Wang, Q. Wu, J. Yang, K. Dong, X. Chen, X. Bai, X. Chen, Z. Chen, C. Viboud, M. Ajelli et al., “Global, regional, and national estimates of target population sizes for covid-19 vaccination: descriptive study,” bmj, vol. 371, 2020.

[12] https://www.who.int/who-documents-detail/draf-landscape-of-covid-19-candidate-vaccines.

[13] G. Giordano, M. Colaneri, A. Di Filippo, F. Blanchini, P. Bolzern, G. De Nicolao, P. Sacchi, P. Colaneri, and R. Bruno, “Modeling vaccination rollouts, sars-cov-2 variants and the requirement for non-pharmaceutical interventions in italy,” Nature Medicine, vol. 27, no. 6, pp. 993–998, 2021.

[14] https://mohfw.gov.in.

[15] J. Yang, V. Marziano, X. Deng, G. Guzzetta, J. Zhang, F. Trentini, J. Cai, P. Poletti, W. Zheng, W. Wang et al., “Can a covid-19 vaccination program guarantee the return to a pre-pandemic lifestyle?” medRxiv, 2021.

[16] K. M. Bubar, K. Reinholt, S. M. Kissler, M. Lipsitch, S. Cobey, Y. H. Grad, and D. B. Larremore, “Model-informed covid-19 vaccine prioritization strategies by age and serostatus,” Science, vol. 371, no. 6532, pp. 916–921, 2021.

[17] G. Béraud, “Mathematical models and vaccination strategies,” Vaccine, vol. 36, no. 36, pp. 5366–5372, 2018.

[18] W. O. Kermack and A. G. McKendrick, “A contribution to the mathematical theory of epidemics,” Proceedings of the royal society of london. Series A, Containing papers of a mathematical and physical character, vol. 115, no. 772, pp. 700–721, 1927.

[19] V. Piccirillo, “Nonlinear control of infection spread based on a deterministic seir model,” Chaos, Solitons & Fractals, vol. 149, p. 111051, 2021.

[20] K. Sarkar, S. Khajanchi, and J. J. Nieto, “Modeling and forecasting the covid-19 pandemic in india,” Chaos, Solitons & Fractals, vol. 139, p. 110049, 2020.

[21] P. Samui, J. Mondal, and S. Khajanchi, “A mathematical model for covid-19 transmission dynamics with a case study of india,” Chaos, Solitons & Fractals, vol. 140, p. 110173, 2020.

[22] D. K. Bagal, A. Rath, A. Barua, and D. Patnaik, “Estimating the parameters of susceptible-infected-recovered model of covid-19 cases in india during lockdown periods,” Chaos, Solitons & Fractals, vol. 140, p. 110154, 2020.

[23] G. Pandey, P. Chaudhary, R. Gupta, and S. Pal, “Seir and regression model based covid-19 outbreak predictions in india,” arXiv preprint 2004.00958, 2020.

[24] K. Chatterjee, K. Chatterjee, A. Kumar, and S. Shankar, “Healthcare impact of covid-19 epidemic in india: A stochastic mathematical model,” Medical Journal Armed Forces India, vol. 76, no. 2, pp. 147–155, 2020.

[25] T. Oraby, M. G. Tyshenko, J. C. Maldonado, K. Vatcheva, S. Elsaadany, W. Q. Alali, J. C. Longenecker, and M. Al-Zoughool, “Modeling the effect of lockdown timing as a covid-19 control measure in countries with differing social contacts,” Scientific reports, vol. 11, no. 1, pp. 1–13, 2021.

[26] A. Senapati, S. Rana, T. Das, and J. Chattopadhyay, “Impact of intervention on the spread of covid-19 in india: A model based study,” Journal of Theoretical Biology, vol. 523, p. 110711, 2021.

[27] A. Mahajan, N. A. Sivadas, and R. Solanki, “An epidemic model sipherd and its application for prediction of the spread of covid-19 infection in india,” Chaos, Solitons Fractals, vol. 140, p. 110156, 2020. [Online]. Available: https://www.sciencedirect.com/science/article/pii/S096007792030552X

[28] A. Mahajan, R. Solanki, and N. Sivadas, “Estimation of undetected symptomatic and asymptomatic cases of covid-19 infection and prediction of its spread in the usa,” Journal of medical virology, vol. 93, no. 5, pp. 3202–3210, 2021.

[29] https://www.worldometers.info/coronavirus.

[30] https://ourworldindata.org/grapher/full-list-covid-19-tests-per-day.

[31] https://www.covid19india.org.

